# Age-dependence of healthcare interventions for COVID-19 in Ontario, Canada

**DOI:** 10.1101/2020.09.01.20186395

**Authors:** Irena Papst, Michael Li, David Champredon, Benjamin M. Bolker, Jonathan Dushoff, David J.D. Earn

**Affiliations:** Center for Applied Mathematics, Cornell University; Department of Biology, McMaster University; South African Centre for Epidemiological Modelling and Analysis, University of Stellenbosch; Department of Pathology and Laboratory Medicine, Western University; Michael G. DeGroote Institute for Infectious Disease Research, McMaster University; Department of Mathematics & Statistics, McMaster University; Department of Mathematics, University of Toronto

**Keywords:** epidemiology, infectious disease, SARS-CoV-2, COVID-19, age distribution, hospitalization

## Abstract

**Background:** Patient age is the most salient clinical indicator of risk from COVID-19. Age-specific distributions of known SARS-CoV-2 infections and COVID-19-related deaths are available for many regions. Less attention has been given to the age distributions of serious medical interventions administered to COVID-19 patients, which could reveal sources of potential pressure on the healthcare system should SARS-CoV-2 prevalence increase.

**Methods:** We analysed 97,957 known SARS-CoV-2 infection records for Ontario, Canada, from 23 January 2020 to 26 November 2020 and estimated the age distributions of hospitalizations, Intensive Care Unit admissions, intubations, and ventilations. We quantified the probability of hospitalization given known SARS-CoV-2 infection, and of survival given COVID-19-related hospitalization.

**Results:** The distribution of hospitalizations peaks with a wide plateau covering ages 54–90, whereas deaths are concentrated in very old ages. The estimated probability of hospitalization given known infection reaches a maximum of 30.9% at age 80 (95% CI 28.0%–33.9%). The probability of survival given hospitalization is near 100% for adults younger than 40, but declines substantially after this age; for example, a hospitalized 54-year-old patient has a 91.5% chance of surviving COVID-19 (95% CI 87.0%–94.9%).

**Conclusions:** Ontario’s healthcare system has not been overstretched by COVID-19 thanks to wide-spread infection control efforts, yet the probability of survival given hospitalization for COVID-19 is lower than is generally perceived for patients over 40. As prevalence continues to increase during this most recent wave of infection, healthcare capacities are at risk of being exceeded. Survival of individuals in the broad age range requiring acute care could decrease, potentially expanding the distribution of COVID-19-related deaths toward younger ages.

## Background

Severe acute respiratory syndrome coronavirus 2 (SARS-CoV-2) was first confirmed in Ontario, Canada, on 23 January 2020, in an individual with a recent travel history to Wuhan, Hubei Province, China [1], the site of the first large-scale outbreak of this novel pathogen [2]. The virus was detected sporadically in Ontario through February [3], until the number of known infections (KIs) began to rise consistently in March. The province declared a state of emergency on 17 March 2020 [4], implementing a large-scale economic shutdown and school closures to help mitigate the spread of the virus. The province began reopening in stages through the summer [5] amid relatively low SARS-CoV-2 infection prevalence, with most schools reopening early-to-mid September [6]. In late September, additional restrictions were enacted across the province in response to the beginning of the second wave of infection [7, 8, 9], and on 23 November 2020 a lockdown was enacted in the densely populated Toronto and Peel regions [10].

A major concern associated with the spread of SARS-CoV-2 is the potential for hospitals to become overwhelmed with COVID-19, the disease caused by the virus, which would compromise care of these patients, and reduce access to and quality of care for many other illnesses. Severity of COVID-19 presentation is highly variable among individuals, but is thought to increase with age [11, 12, 13]. Deaths attributed to COVID-19 have been found to be strongly concentrated in the elderly [14, 15, 16]. Comparatively few studies have explored the age distribution of serious medical interventions administered to COVID-19 patients [17, 18].

## Methods

In order to understand the potential for the healthcare system to become overwhelmed by SARS-CoV-2 infections, it is important to identify the demographic groups that are most likely to require significant medical care. The goal of this study is to quantify the relationships between COVID-19 patient age and the administration of serious medical interventions (hospitalizations, intensive care unit (ICU) admissions, intubations, and ventilations) for the province of Ontario. We compare these age-intervention associations with the age distributions of KIs and deaths. We also estimate the age-specific probability of hospitalization given known SARS-CoV-2 infection, and of survival given hospitalization related to COVID-19.

We use individual-level line lists for known SARS-CoV-2 infections reported up to 26 November 2020 from the Case and Contact Management (CCM) database maintained by Public Health Ontario. This central database includes detailed records of SARS-CoV-2 infections across the entire province of Ontario. These records include an individual’s demographic information (such as age), whether COVID-19-related interventions (including hospitalization, ICU admission, intubation, ventilation) were administered, as well as whether the infection was fatal.

The CCM database includes 111,210 KI records up to 26 November 2020. However, we analyse only KIs marked as “resolved” or “fatal” (*N* = 97,957) to avoid tallying patients whose final outcomes are not yet known. KIs are marked as “resolved” in CCM based on public health unit assessment. In all instances, a record is considered resolved if it is 14 days past the symptom onset date (or specimen collection if symptom onset date is not known), though public health occasionally performs additional follow-up to update records. For brevity, we use “resolved KIs” in our analysis to refer to KIs marked as either “resolved” or “fatal” in CCM.

These data are representative of a situation where the healthcare system was not overwhelmed with COVID-19 patients (due to wide-spread measures to keep SARS-CoV-2 spread under control). ICU occupancy for COVID-19 treatment in Ontario never exceeded more than 13% of the total ICU capacity over the period covered by the data. We calculate this percentage using daily ICU occupancy from publicly-available data [19] and the number of ICU (critical care) beds available according to the Government of Ontario [20]. Based on the latter source, we assume the province had a total of 2012 ICU beds before 16 April 2020, and this capacity increased to 3504 beds on 16 April 2020. We acknowledge that this is a simplification of the ICU expansion effort and that not all ICU beds are allocated to COVID-19 treatment (although this allocation is flexible depending on demand).

For population counts, we use 2020 Ontario population projections produced by Statistics Canada [21], specifically projections from the “M1” medium-growth scenario [22], although all scenarios yield virtually identical projections for the short time horizon of 2020.

We use provincial SARS-CoV-2 testing data from the Ontario Laboratories Information System (OLIS) database. This database records all tests for active SARS-CoV-2 infection in the province. These data are based on reports up to 26 November 2020 and include 90,853 positive tests and 2,957,327 negative tests.

We aggregate counts of all age-specific data (KIs, population, and tests) into two-year age bins, with one wide bin for individuals over 100 years old.

We use a binomial generalized additive model to estimate the age-specific probability of hospitalization given KI [23], and a binomial generalized linear model to estimate the age-specific survival probability given hospitalization [24].

## Results

Figure 1 shows the time course of the SARS-CoV-2 epidemic in Ontario up to 26 November 2020, as represented by KIs.

**Figure 1:**
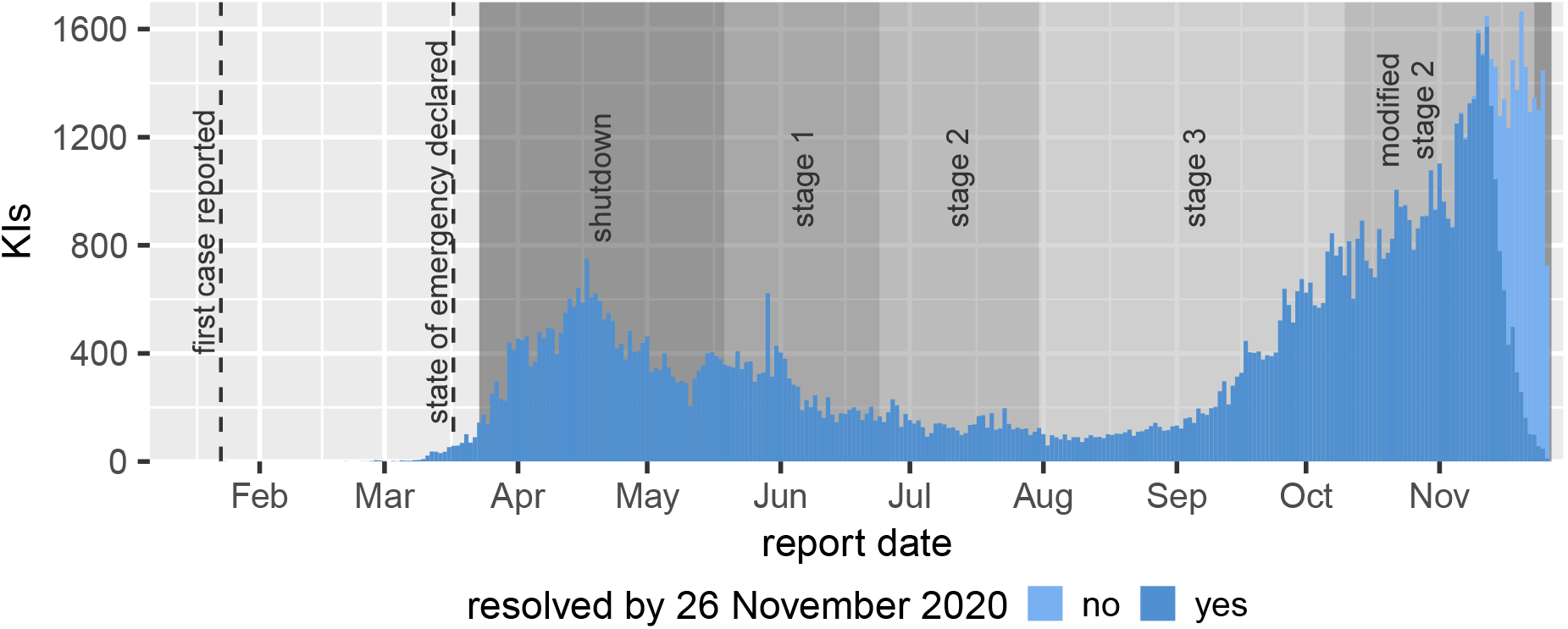
Known infections (KIs) over time in Ontario. Counts are split by whether or not the KI was resolved (marked as “resolved” or “fatal” in the Case and Contact Management database) by 26 November 2020 (see Methods). Dashed vertical lines mark important dates for the outbreak in the province. Shaded regions indicate roughly when the most densely populated regions were in each reopening stage (reopenings did not occur uniformly across public health units). Descriptions of each reopening stage can be found on the official Ontario COVID-19 website [5].

Figure 2 shows the age structure of resolved KIs, population demographics, and SARS-CoV-2 infection tests. The pattern observed in the raw counts of resolved KIs (panel A) reflects underlying demographics (panel B) as well as the count of positive tests per capita, which is in turn related to testing intensity (panel D) Testing intensity is defined as the number of tests administered per 10,000 population by age group. Controlling for demography, the number of detected infections (panel C) is relatively low in ages under 15, increases to a relative plateau for ages 20–70 (with a noticable crest around age 25), and then continually increases after age 70. The number of resolved KIs per capita is 1.45 times higher in ages 20–29 than in ages 30–69. Testing intensity increases after age 75.

**Figure 2:**
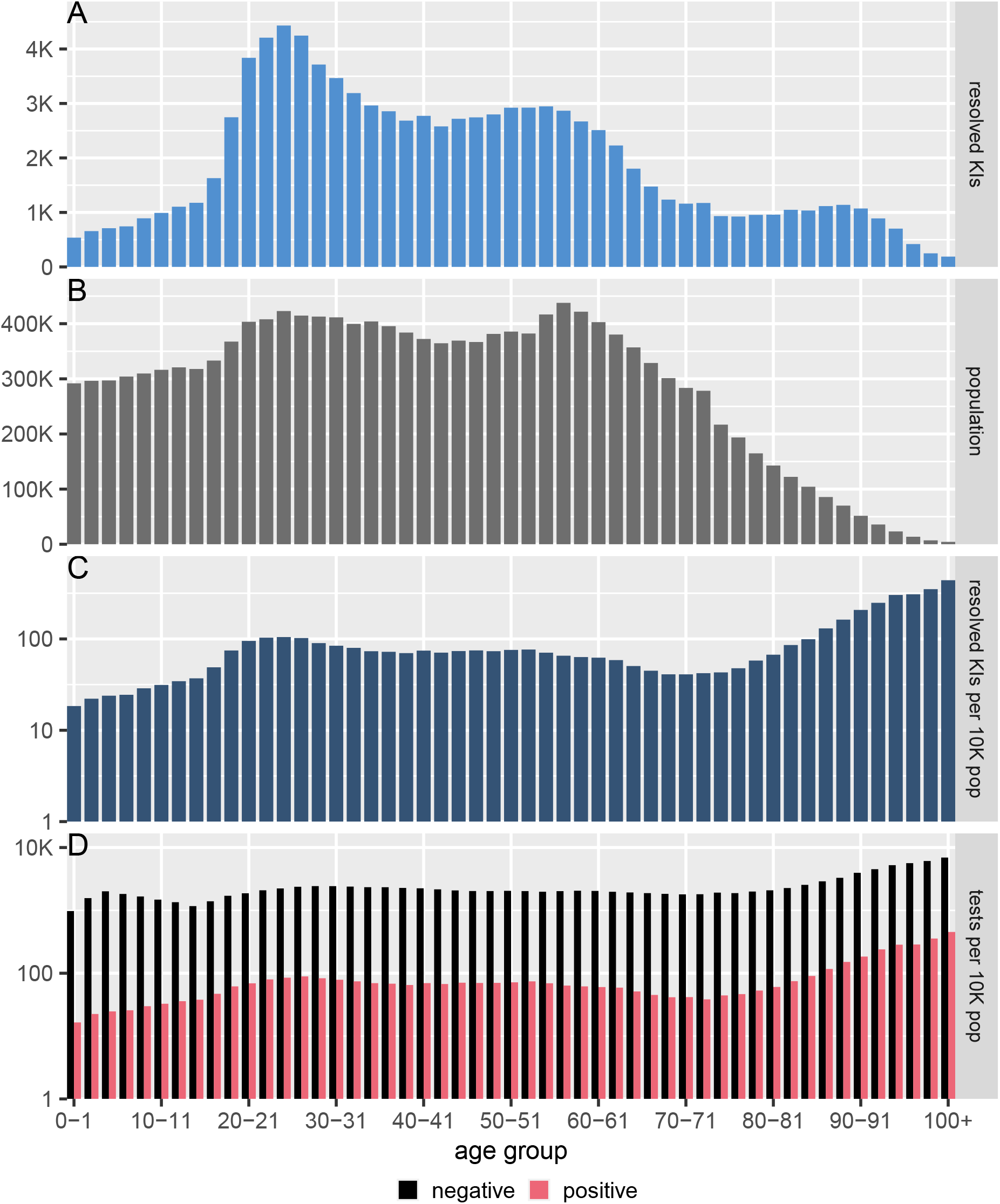
Age distribution of known infections (KIs) in Ontario. The distribution of ages for resolved KIs (panel A), Ontario population projections for 2020 (panel B), resolved KIs per 10,000 population (panel C), and SARS-CoV-2 infection tests per 10,000 population (panel D). The y-axes in panels C and D are on a logarithmic scale.

Figure 3 shows the distribution of ages for serious medical interventions (panel A) and deaths (panel B) related to COVID-19 for resolved KIs. We present raw counts, as opposed to counts normalized by the age-specific population, because the counts (not per capita counts) determine the pressure on the healthcare system. Hospitalizations are faceted by the most intensive known intervention (with ventilator use being the most intensive, followed by intubation, then ICU admission, and hospitalization). Deaths are split by whether or not the patient has a record of hospitalization for COVID-19 treatment.

**Figure 3:**
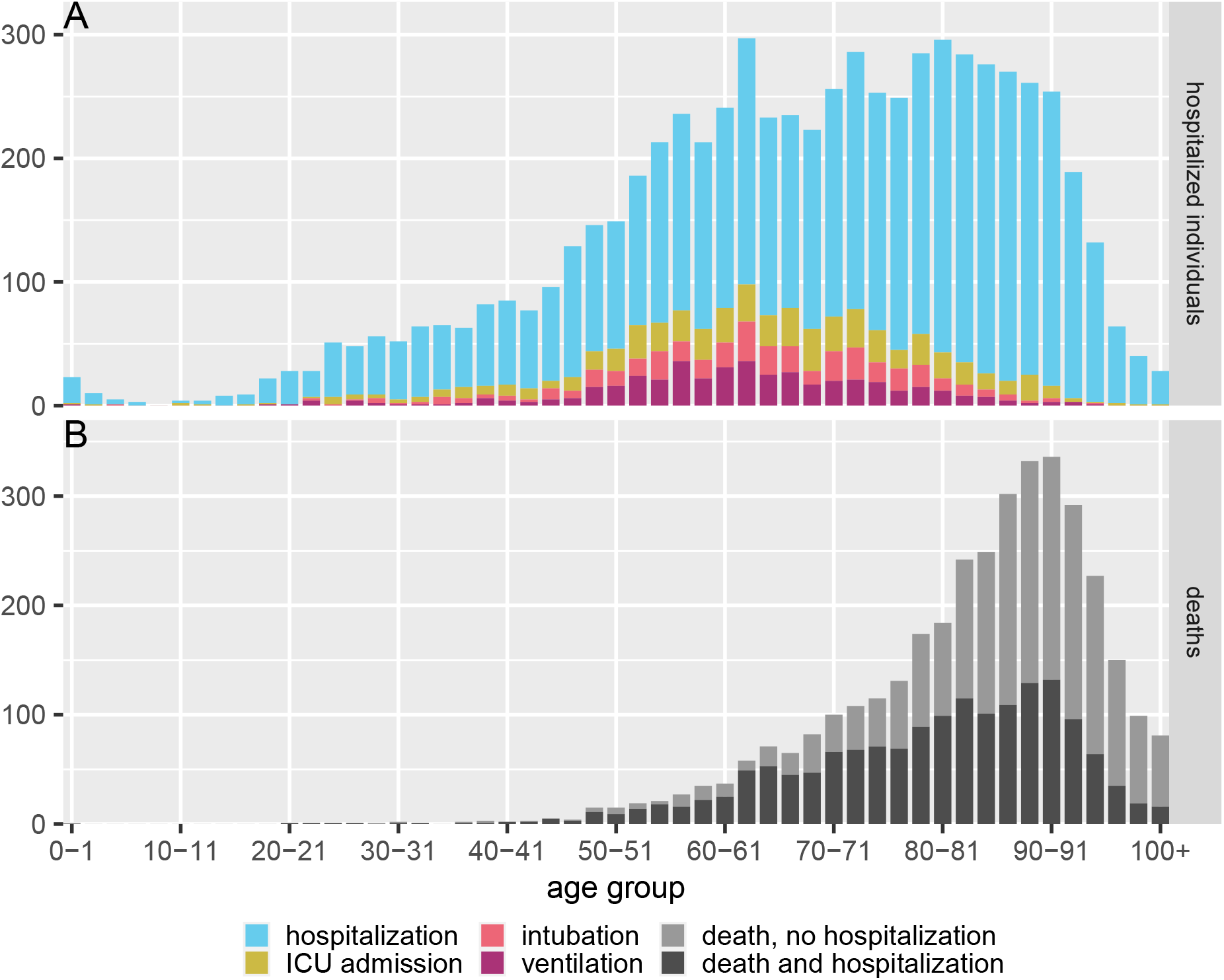
COVID-19 outcomes by age in Ontario. The distribution of ages for hospital interventions (panel A), and deaths (panel B). Hospital outcomes are nested and tallied by the most intensive medical intervention used for each patient (ventilator use is the most intensive, followed by intubation, ICU admission, and hospitalization). Deaths are faceted by whether or not the patient also had a record of hospitalization for COVID-19 treatment.

The distribution of serious medical interventions is much wider than that of deaths, with the latter peaking at age 90. Hospitalizations are relatively uniformly spread between ages 54–90, while the distribution of ICU-related interventions (ICU admission, intubation, ventilation) is spread over a slightly younger age range. The majority of deaths, 55.3%, have occurred in KIs where there is no record of hospitalization for treatment related to COVID-19.

Figure 4 shows the estimated hospitalization probability given known SARS-CoV-2 infection (panel A) and survival probability given hospitalization for COVID-19 treatment (panel B). The large uncertainty in these probability estimates for some young and very old age groups is due to small numbers of KIs and hospitalizations in these ages. The hospitalization probability peaks in the 80–81 age group at 30.9% (95% CI 28.0%–33.9%). In adults, the survival probability is near 100% until about age 40, where it begins to decline steadily. For instance, a hospitalized individual in the age group 54–55 has a 91.5% chance of surviving COVID-19 (95% CI 87.0%–94.9%), implying that nearly 1 in 10 hospitalized COVID-19 patients in this age group die despite receiving medical care while ICUs and hospitals are below capacity.

**Figure 4:**
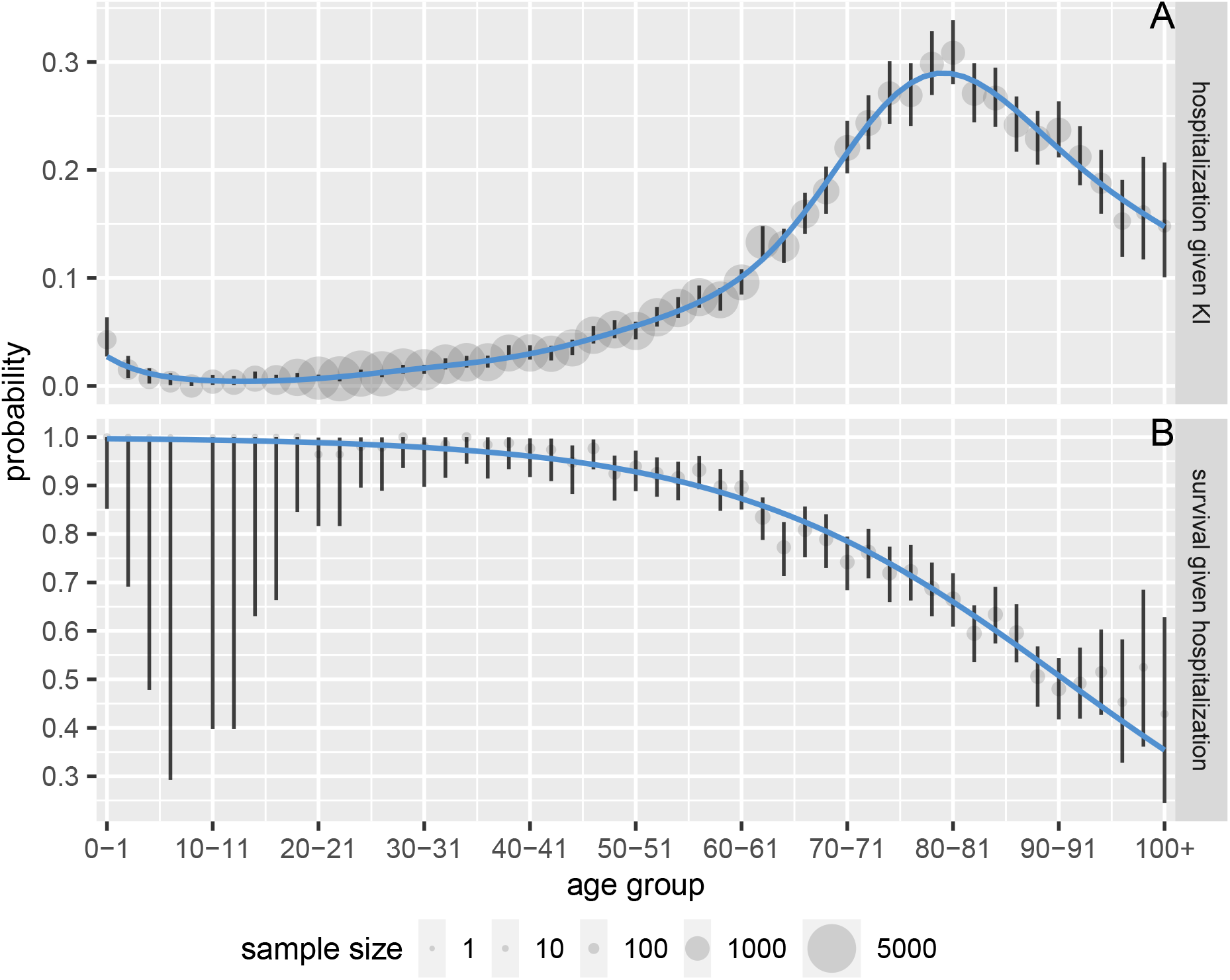
Age-dependent COVID-19 hospitalization probability for known SARS-CoV-2 infection (panel A) and survival probability for hospitalized patients (panel B) in Ontario. We estimate the hospitalization probability using a generalized additive model and the survival probability using a generalized linear model (blue curves; see Methods). Vertical lines give age-specific 95% binomial confidence intervals and point areas are proportional to samples sizes.

## Discussion

The age distribution of known SARS-CoV-2 infections (Figure 2A) and deaths attributed to COVID-19 (Figure 3B) alone provide limited insight into the risk that COVID-19 patients could overwhelm Ontario’s healthcare system. The majority of COVID-19-related deaths have occurred in patients with no record of hospitalization (Figure 3B), and many deaths to date have occurred in long-term care homes, which are independent of the hospital system [25]. The broad age distributions of hospitalizations, ICU admissions, intubation, and ventilation (Figure 3A) reveal the potential pressure on the healthcare system from both middle-aged individuals and seniors.

Early in the outbreak, the province of Ontario expanded coverage for COVID-19-related treatment to include even individuals who are not usually covered by the Ontario Health Insurance Plan [26]. Access to prompt and successful medical interventions may have kept a large proportion of COVID-19-related hospitalizations from resulting in deaths. While we expect that the shape of the age distribution of the *need* for hospitalization (Figure 3A) would remain the same if prevalence were to increase further (as is occurring at the time of writing), the distribution of deaths (Figure 3B) may expand toward younger ages if hospitals and ICUs reach maximum capacity.

Relatively high rates of resolved KIs and testing (Figure 2C-D) in those over age 75 can be partly attributed to significant outbreaks and disease surveillance in long-term care facilities [25, 27].

The age-dependent probabilities of hospitalization given KI (Figure 4A) are based on resolved *known* infections, and so they depend on how widely SARS-CoV-2 testing has been conducted. Throughout the period covering a large portion of the CCM data, testing guidelines selected for sufficiently symptomatic individuals [28]. These guidelines were not expanded to include asymptomatic individuals from the general public until 29 May 2020 [29] and were rolled back on 24 September 2020 in an effort to preserve limited testing resources amid a surge in KIs [30]. As a result, untargetted asymptomatic testing was offered only in the summer, when prevalence was relatively low, which represents a small proportion of the data. Moreover, individuals may not be prompted to get tested in the absence of symptoms unless they are included in a contact tracing investigation. The probability of hospitalization given KI therefore likely overestimates the underlying probability of hospitalization given infection, whether known or not.

Our survival probability estimates for hospitalized individuals (Figure 4B) are *not* affected by the same detection biases present in KI data. Patients admitted to hospital are tested for SARS-CoV-2 as part of infection control protocols, and thus infection detection in hospitalized individuals is not influenced by testing guidelines for the general population. Our survival probability estimates do, however, represent an upper bound with respect to the current standard of care and viral variant. In the absence of significant innovation in COVID-19 treatment or viral evolution to lower disease severity, we expect survival probabilities would decrease if ICUs or hospitals were to reach maximum capacity.

## Limitations

In general, the number of KIs underestimates the true prevalence of SARS-CoV-2 infection for a variety of reasons, including test availability, ease of testing, test accuracy, and difficulties in detecting asymptomatic individuals. The majority of known infections captured in the data analysed in this study occurred when testing guidelines were selecting for sufficiently symptomatic individuals, and so asymptomatic and mild infections are likely underrepresented. Contact patterns in Ontario have changed over the course of the pandemic due to the province’s continuing effort to control COVID-19 spread while also supporting the economy. Observed patterns in the age distributions of KIs and deaths may change as the age-specific contact structure and contact rates continue to change.

### Conclusions

We have quantified the age distributions of serious medical interventions for SARS-CoV-2 infection in Ontario, Canada, for the entire period of the regional epidemic. Wide-spread infection control efforts have kept Ontario’s healthcare system from being overstretched, yet the probability of survival given COVID-19-related hospitalization is lower than is generally perceived for patients over 40. Ontario is currently experiencing a significant increase in prevalence of COVID-19; if healthcare capacities were to be exceeded, the distribution of COVID-19-related deaths might expand toward younger ages, where there is already a notable demand for acute care.

The Government of Canada and the Province of Ontario have implemented policies meant to help mitigate SARS-CoV-2 spread while also undertaking a phased reopening [31]. Future work should consider whether the age dependence of SARS-CoV-2 infection risks is changing over time, as the population continues to navigate the pandemic and the testing effort expands. Our study explores only short-term SARS-CoV-2 infection outcomes; future studies should explore the age distributions of long-term morbidities from this infection, so that we may better understand the heterogeneous risks associated with COVID-19. Lastly, all studies relying on known infection counts are subject to bias from how infections are detected via active infection testing. Future work should seek to correct for this bias.

## Data Availability

Data and source code for all analyses will be provided in an Open Science Framework repository upon publication of this manuscript.

## Acknowledgements

We thank Sarah Morrison for assistance with literature review. We are grateful to the Ontario COVID-19 Modelling Table and Science Table (https://covid19-sciencetable.ca/) for valuable discussions. Data were kindly provided by Public Health Ontario. We thank ICES for providing access to data used for preliminary analyses.

This study was supported by the Ontario Health Data Platform (OHDP), a Province of Ontario initiative to support Ontario’s ongoing response to COVID-19 and its related impacts. The opinions, results and conclusions reported in this paper are those of the authors and are independent from the funding sources. No endorsement by the OHDP, its partners, or the Province of Ontario is intended or should be inferred.

## Funding

DJDE, BMB, and JD were funded by the Natural Sciences and Engineering Research Council of Canada, the Michael G. DeGroote Institute for Infectious Disease Research at McMaster University, and the Public Health Agency of Canada.

## Abbreviations

SARS-CoV-2: Severe Acute Respiratory Syndrome Coronavirus 2
KI: Known Infection
COVID-19: Coronavirus Disease 2019
ICU: Intensive Care Unit
CCM: Case and Contact Management
OLIS: Ontario Laboratories Information System

## Ethics approval and consent to participate

The study received ethics approval from the Health Sciences Research Ethics Board at the University of Toronto.

## Competing interests

The authors declare that they have no competing interests.

## Consent for publication

Not applicable.

## Authors’ contributions

IP and DJDE designed this research project, performed the data analysis, and drafted the manuscript. BMB provided specific guidance for statistical analyses. All authors discussed and interpreted the results, revised the manuscript, and approved the final version for publication.

## Notes

### Competing Interest Statement

The authors have declared no competing interest.

### Summary of Updates

Updated data and revised text to reflect the current situation in Ontario.

